# Mining the Health Disparities and Minority Health Bibliome: A Computational Scoping Review and Gap Analysis of 200,000+ Articles

**DOI:** 10.1101/2023.10.17.23296754

**Authors:** Harry Reyes Nieva, Suzanne Bakken, Noémie Elhadad

## Abstract

Without comprehensive examination of available literature on health disparities and minority health (HDMH), the field is left vulnerable to disproportionately focus on specific populations or conditions, curtailing our ability to fully advance health equity. Using scalable open-source methods, we conducted a computational scoping review of more than 200,000 articles to investigate major populations, conditions, and themes in the literature as well as notable gaps. We also compared trends in studied conditions to their relative prevalence in the general population using insurance claims (42MM Americans). HDMH publications represent 1% of articles in MEDLINE. Most studies are observational in nature, though randomized trial reporting has increased five-fold in the last twenty years. Half of all HDMH articles concentrate on only three disease groups (cancer, mental health, endocrine/metabolic disorders), while hearing, vision, and skin-related conditions are among the least well represented despite substantial prevalence. To support further investigation, we also present HDMH Monitor, an interactive dashboard and repository generated from the HDMH bibliome.

## 1. Introduction

In 1985, the Department of Health and Human Services (DHHS) *Report of the Secretary’s Task Force on Black and Minority Health* characterized the legacy of disparities in healthcare and health outcomes among racial and ethnic groups in the United States “as an affront both to our ideals and the ongoing genius of American medicine” (1). Commonly known as the Heckler Report, this landmark document garnered unprecedented national attention for health disparities and minority health (HDMH). Over the years, additional evidence such as *Unequal Treatment: Confronting Racial and Ethnic Disparities in Health Care* from the Institute of Medicine (now the National Academy of Medicine) have supported further inquiry to elucidate minority health and advance health equity (2). Such efforts culminated in the formation and mandate of the National Institute on Minority Health and Health Disparities (NIMHD).

Over the last half-century, HDMH researchers have generated a vast amount of scientific evidence and developed important interventions. During this same period, data availability, quality, and the capacity to leverage it have dramatically improved. Our understanding of the basic mechanisms and etiology of disease and potential avenues for intervention have also advanced. Despite substantial concerted efforts, however, reduction of health disparities remains limited and the unique health needs of minority populations remain to be fully characterized (3).

To transform the HDMH research landscape and promote a new paradigm for the advancement of health disparities science, the NIMHD recently embarked on a science visioning process (4). This work identified thirty strategies divided among three pillars – methods and measurement, etiology, and interventions – to foster a new generation of research capable of making major strides to improve minority health and reduce health disparities. Among the strategies proposed, the NIMHD promotes standardized rigorous methods and measurements that ensure health disparities research is relevant to a diversity of populations and leverages multiple sources of existing and emerging data to enhance analytical capacity (5, 6).

Given the considerable amount of scientific knowledge available to clinicians, researchers, and policy makers, a comprehensive examination of available HDMH literature may further elucidate important insights and inform future research planning. Highly specialized systematic and narrative reviews concerning HDMH have been conducted (6–20). Nonetheless, due to limitations of scope and scale associated with manual assessment, a comprehensive scoping review of the HDMH literature has not been performed, leaving the field potentially vulnerable to disproportionately focus on specific populations or emphasize certain conditions, curtailing our ability to fully advance health equity and improve our understanding of the health of minority populations.

Computational approaches, natural language processing (NLP) and machine learning (ML) in particular, have proven capable of surmounting traditional constraints on reviews using reproducible, scalable methods (21–23). The objective of this study was to perform a comprehensive, computational scoping review of the HDMH bibliome to investigate the following questions:

1. What are the major populations, study methods, conditions, and themes in the HDMH literature?
2. How have dominant themes changed over time?
3. What gaps exist in the literature?

We also make our findings publicly available via HDMH Monitor, a new interactive dashboard and article repository (https://hdmhmonitor.dbmi.columbia.edu), to support future scientific discovery via automated search and archive of HDMH articles, information synthesis, and data visualization.

## 2. Results

Guided by the MEDLINE/PubMed Health Disparities and Minority Health Search Strategy (24) established by the National Library of Medicine (NLM), the final scoping review corpus contained 206,011 articles (**Figure S1**). This pre-validated query incorporates 257 subject terms in addition to searching publication titles and abstracts to identify articles relevant to HDMH (**Table S1)**. Search terms include, for example, “health inequities”, “social determinants of health”, “African Americans”, and “undocumented immigrants.” The search parameters do not include any disease-specific terms that might bias our results. MEDLINE and its public search interface, PubMed, are the primary tools for performing biomedical literature searches among health professionals and students (25–28). Large-scale analyses revealed notable trends and gaps in the HDMH literature. Results presented are based on examination of Medical Subject Headings (MeSH) assigned to articles in MEDLINE, conditions mentioned in abstracts and titles, emerging themes derived from the HDMH literature, and comparison of condition coverage in the literature and prevalence in national insurance claims data. A graphical abstract summarizing our approach is provided in **Figure 1**.

**Figure 1:**
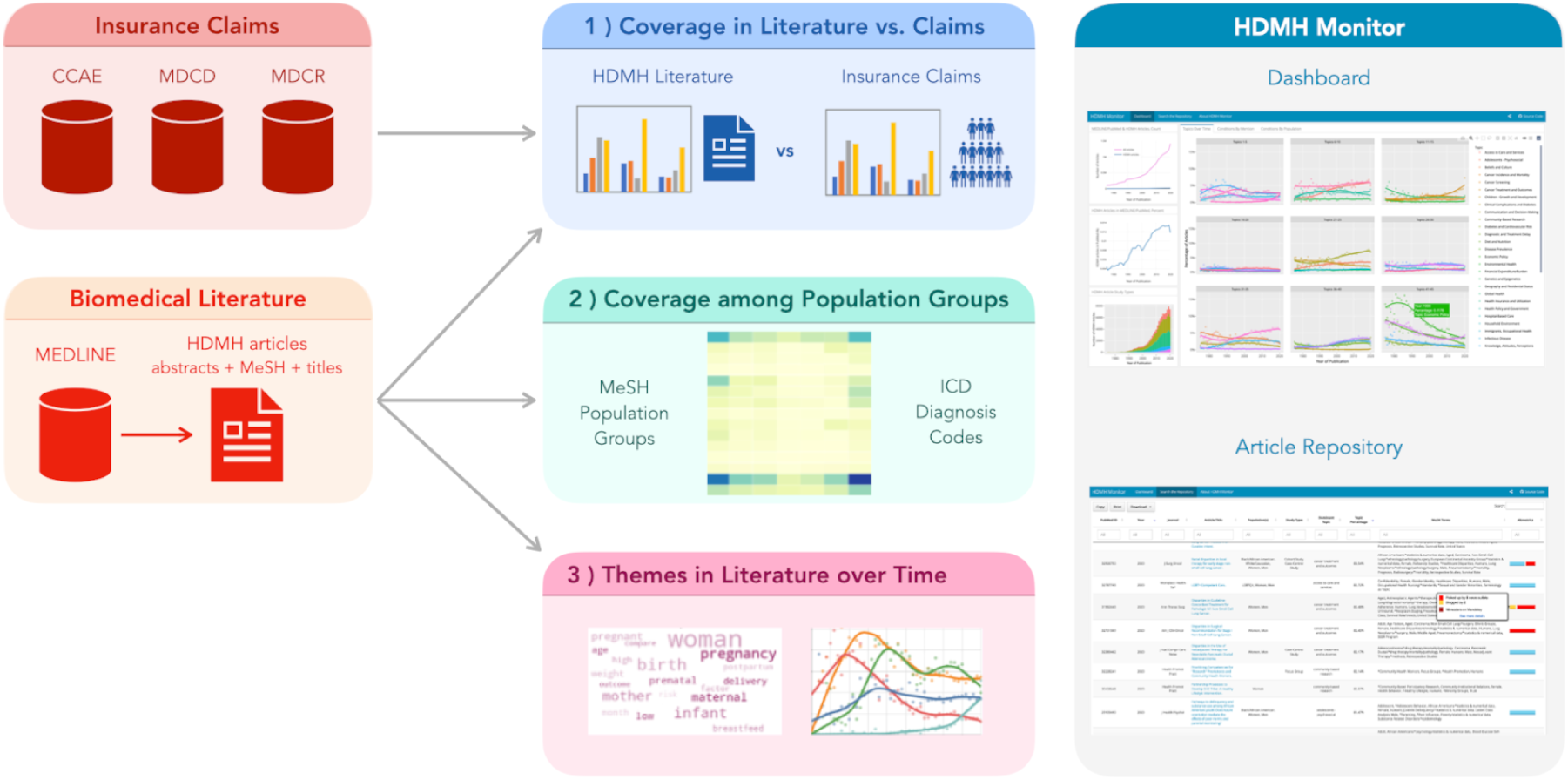
Graphical abstract. We identified HDMH articles using the MEDLINE/PubMed Health Disparities and Minority Health Search Strategy and indexed them via MeSH terms, conditions mentioned, and themes derived from topic modeling. We also compared trends in studied conditions to their relative prevalence in the general population using data from external insurance claims databases. *Abbreviations:* CCAE, Commercial Claims And Encounters; HDMH, Health Disparities and Minority Health; MDCD, Medicaid; MDCR, Medicare; MEDLINE, Medical Literature Analysis and Retrieval System Online; MeSH, Medical Subject Headings.

### 2.1 Production of HDMH scientific publications has increased but is largely limited to observational studies and traditional analytical methods

We examined patterns in production of scientific articles from 1975, a decade prior to release of the landmark Heckler Report, through 2020 as the COVID-19 pandemic dominated the scientific and global landscape. HDMH articles comprised 0.5% (408 articles) of MEDLINE references published in 1975 compared to approximately 1% (13,180 articles) in 2020 (**Figure 2**). HDMH article production was relatively consistent from 1975 through the mid-1980s (0̃.5% of MEDLINE each year), then began to increase sharply before stabilizing during the majority of the 1990s. Notably, this rise in HDMH publications occurs just after the Heckler Report brought unprecedented national attention to health disparities. Similarly, production started to rebound after an initial drop fifteen years later in 2000 following the formation of the National Center on Minority Health and Health Disparities (NCMHD), later renamed the NIMHD. In 2020, we note a sharp decrease in production as a percentage of MEDLINE. While this decline is striking, this same year also exhibited the highest annual count of articles found in MEDLINE in a single year (both overall and among HDMH articles). The increase in publication output has been largely attributed to the COVID-19 pandemic (29–31). It has also been asserted that, as most journals did not increase the total number of articles published per issue, the proliferation of COVID-related articles occurred at the expense of non-COVID-related publications (31). Nonetheless, many articles also served to spotlight the numerous disparities illustrated by the pandemic (32). Our corpus of HDMH literature contained 1,205 articles related to COVID-19 (8% of all HDMH articles published in 2020).

**Figure 2:**
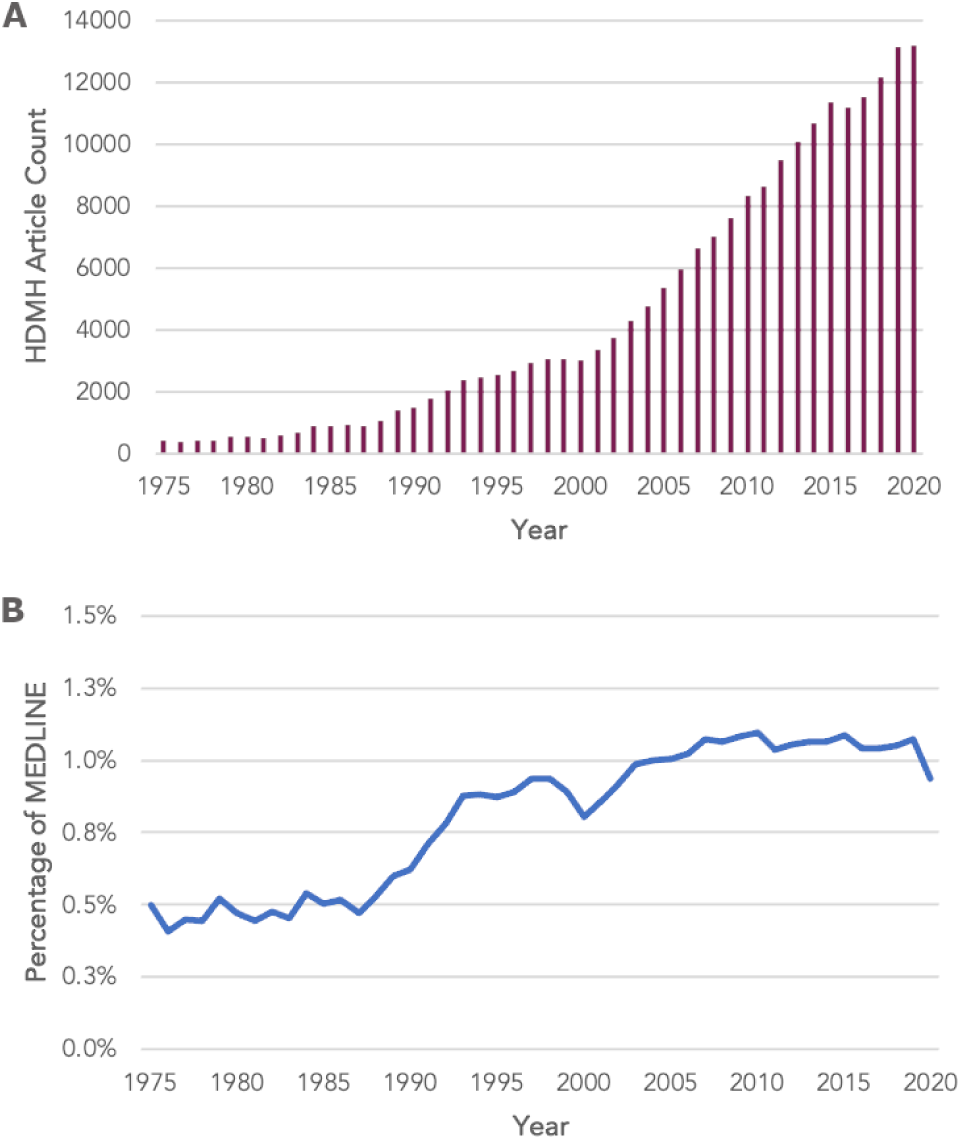
Health Disparities and Minority Health (HDMH) Publications (N=206,011) by (A) Percentage of MEDLINE and (B) Article Count. Between 1975 and 2020, HDMH research publications in MEDLINE, the National Library of Medicine bibliographic database containing more than 28 million biomedical references from over 5,200 journals, rose annually from 0.5% (408 articles) to 1% (13,180 articles).

Based on review of MeSH terms, we found 7,869 HDMH articles (4% of corpus) reporting on randomized controlled trials (RCTs). From 2001 to 2020, the annual number of articles on RCTs increased five-fold from 102 to 513, respectively. Among all articles in the HDMH corpus, we identified 167 with MeSH tags related to artificial intelligence (e.g., ML, NLP, neural networks, or their descendants in the MeSH hierarchy).

### 2.2 Half of all HDMH articles focus on three disease categories

We mapped conditions mentioned in the titles and abstracts of HDMH articles to the International Classification of Diseases, Tenth Revision (ICD-10), then examined the distribution of articles among ICD-10 chapters, the highest-level grouping for a set of related diagnoses. ICD-10 is routinely used by physicians to code clinical diagnoses for insurance claims. While conditions across all ICD-10 chapters were identified in the literature (**Table 1**), half (49%) of all HDMH articles concentrated on three ICD chapters – II: Neoplasms (17% of all articles), V: Mental, behavioral, and neurodevelopmental disorders (16%), and IV: Endocrine, nutritional, and metabolic diseases (16%).

**Table 1:**
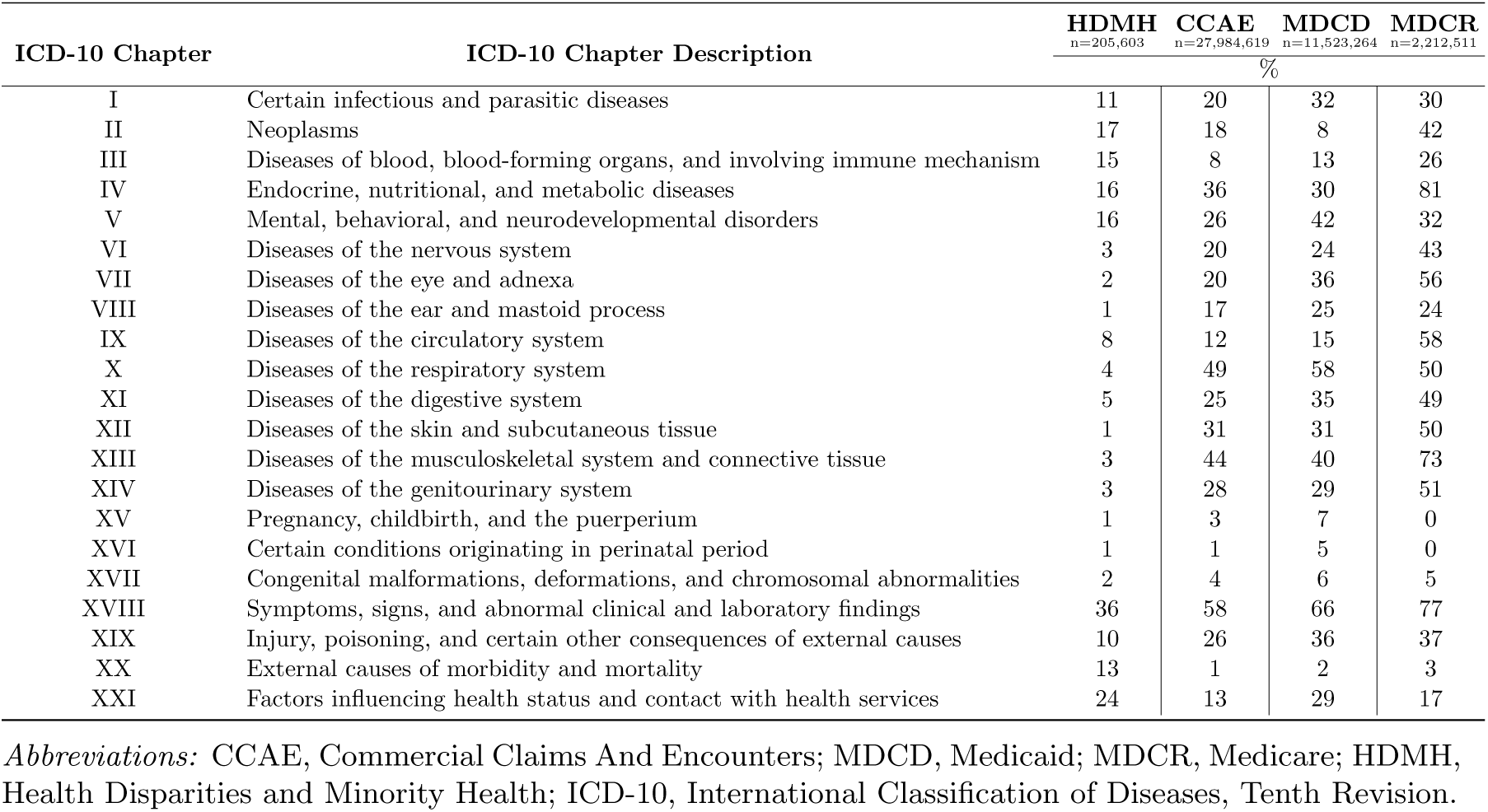
ICD-10 Disease Chapters by Percentage of Relevant Articles in HDMH Literature and Prevalence in MarketScan Claims Databases. Half (49%) of all HDMH articles studied conditions associated with three ICD-10 Chapters (II, IV, V). Upon comparison, coverage in the HDMH literature (n=206,011) differed substantially from clinical prevalence in government and private insurance claims datasets representing nearly 42 million Americans.

Analysis also revealed that nearly a quarter of articles (23%) mentioned either external causes of morbidity and mortality (Chapter XX) or injury, poisoning and certain other consequences of external causes (Chapter XIX). Diseases of blood, blood-forming organs, and involving immune mechanism (Chapter III) comprised 15% of articles, while nearly one in ten HDMH articles referred to certain infectious and parasitic diseases (Chapter I) and diseases of the circulatory system (Chapter IX). Pregnancy, childbirth, and the puerperium (Chapter XV), certain conditions originating in the perinatal period (Chapter XVI), and congenital malformations, deformations, and chromosomal abnormalities (Chapter XVII) were referenced in 4% of all articles. Diseases of the ear and mastoid process (Chapter VIII), diseases of the skin and subcutaneous tissue (Chapter XII), and diseases of the eye and adnexa (Chapter VII) were the least well represented (1%, 1%, and 2% of HDMH articles, respectively).

Symptoms, signs, and abnormal clinical and laboratory findings (Chapter XVIII) and Factors influencing health status and contact with health services (Chapter XXI) lack codes for specific diseases or other diagnoses. Accordingly, while codes from these chapters were commonly found, their detection did not contribute to our primary aim – identifying diseases or other conditions and characterizing the degree to which they are represented in the literature. Nonetheless, we report their values in tables and figures in the interest of completeness and to inform potential future work.

### 2.3 Disease prevalence in the general population is not necessarily indicative of the foci of HDMH research

Upon comparison, coverage of ICD-10 condition chapters in the HDMH literature differed substantially from clinical prevalence among 42 million Americans represented in MarketScan Commercial Claims And Encounters (CCAE), Medicaid (MDCD), and Medicare (MDCR) datasets (**Table 1**). Mention of external causes of morbidity and mortality (Chapter XX) was found in 13% of HDMH references (ranked fifth highest in chapter coverage based on percentage of articles). In contrast, ranging from 1% to 3%, conditions found in this chapter were among the least prevalent in claims databases (ranked lowest prevalence in CCAE and MDCD).

Condition coverage in the literature and clinical prevalence in the general population were most similar for Chapter II: Neoplasms (17% in HDMH corpus and 18% in CCAE dataset) and Chapter III: Diseases of blood, blood-forming organs, and involving immune mechanism (15% in HDMH corpus and 13% in MDCD dataset). While the neoplasms chapter was among the highest ranked in the HDMH bibliome based on condition coverage (i.e., mentioned in the most articles), it ranked 13th highest of 21 chapters in CCAE, 17th in MDCD, and 11th in MDCR based on clinical prevalence. Conversely, diseases of the skin and subcutaneous tissue (Chapter XII), diseases of the eye and adnexa (Chapter VII), and diseases of the ear and mastoid process (Chapter VIII) were least represented in the literature (1% to 2% of articles), but ranked much higher relatively in claims databases, ranging in overall prevalence from 31% to 50% (Chapter XII), 20% to 56% (Chapter VII), and 17% to 25% (Chapter VIII).

Symptoms, signs, and abnormal clinical and laboratory findings (Chapter XVIII) was the most prevalent ICD-10 chapter across all sources, HDMH bibliome and claims datasets alike. Pregnancy, childbirth, and the puerperium (Chapter XV), certain conditions originating in the perinatal period (Chapter XVI), and congenital malformations, deformations, and chromosomal abnormalities (Chapter XVII) were also consistently ranked among the lowest in clinical prevalence and coverage in the literature.

### 2.4 Disease coverage in the literature is highly variable among minority populations

We identified conditions across all ICD-10 chapters among MeSH categories for minority populations but found high variability in condition coverage across groups (**Figure 3**). Symptoms, signs, and abnormal clinical and laboratory findings (Chapter XVIII) were commonly found across populations. References related to race, ethnicity and geographic region of residence (i.e., urban, rural) were the most well represented in the literature while immigrant populations and persons with disabilities were the least well represented. Cancer (Chapter II: Neoplasms) was the most dominant ICD-10 chapter among articles across racial and ethnic MeSH categories. Similarly, mental, behavioral, and neurodevelopmental disorders (Chapter V) as well as endocrine, nutritional, and metabolic diseases (Chapter IV) exhibited higher relative coverage.

**Figure 3:**
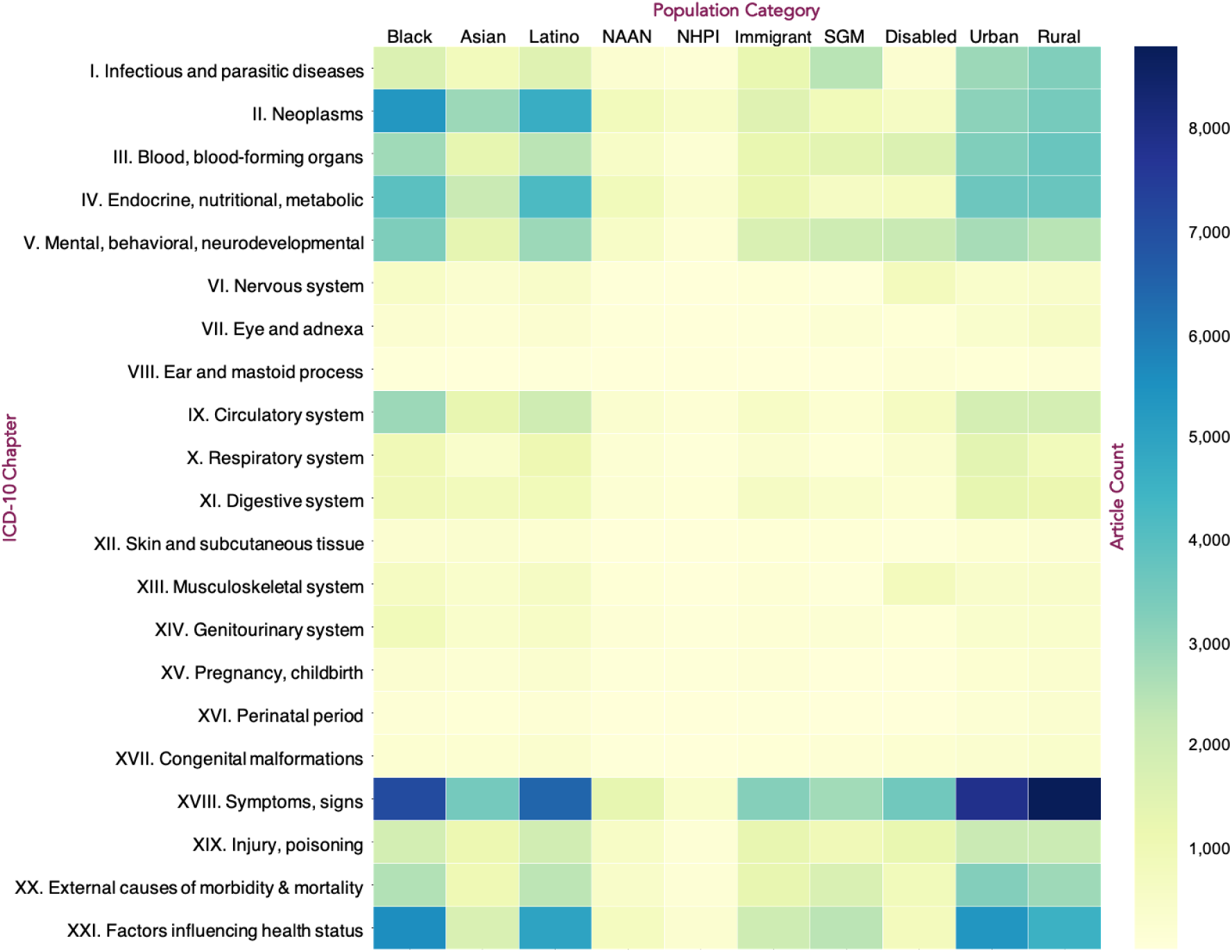
HDMH Article Counts by ICD-10 Chapter and Population Category. Conditions across all ICD-10 chapters (see Table 1 for full descriptions) were identified among minority groups (based on MeSH population categories) with high variability in condition coverage within and across populations. *Abbreviations*: Disabled, Disabled Persons. HDMH, Health Disparities and Minority Health. ICD-10, International Classification of Diseases, Tenth Revision. Latino, Hispanic or Latina/e/o/x. MeSH, Medical Subject Heading. NAAN, Native American or Alaska Native. NHPI, Native Hawaiian or other Pacific Islander. SGM, Sexual and Gender Minorities.

Compared to other minority groups, we noted larger emphasis on diseases of the circulatory system (Chapter IX) among articles tagged with Black/African descent MeSH population categories. Otherwise, ICD-10 chapters exhibited similar trends among people of Asian, Hispanic or Latino, Native American or Alaska Native, and Native Hawaiian or other Pacific Islander descent among relatively fewer articles. In contrast, studies reporting on sexual and gender minorities concentrated on infectious diseases (Chapter I) and mental, behavioral, and neurodevelopmental disorders (Chapter V).

Literature pertaining to immigrant status and persons with disabilities were also represented in the HDMH literature. Articles regarding injury, poisoning, and certain other consequences of external causes (Chapter XIX), external causes of morbidity and mortality (Chapter XX), and mental, behavioral, and neurodevelopmental disorders (Chapter V) were more common than other conditions in both groups. Studies reporting on persons with disabilities also focused on diseases of the musculoskeletal system and connective tissue (Chapter XIII) and diseases of the nervous system (Chapter VI).

### 2.5 Themes in HDMH literature have changed over the last half century

Topic modeling, a computational approach to thematic analysis, revealed themes in the HDMH literature ranging in discussions of groups and subgroups (e.g., racial and ethnic groups, sexual and gender minorities, adolescents, immigrants), conditions (e.g., cancer, cardiovascular disease, diabetes mellitus, infectious disease, overweight and obesity), and study approaches (e.g., survey methods, community-based research, and assessments of knowledge, attitudes, and perceptions). Topic modeling also enabled a nuanced analysis capable of distinguishing between seemingly similar yet distinct themes. For example, maternal-child health (Topic 11) and reproductive health (Topic 33) both share common terms like woman and pregnancy (**Figure 4**). Likewise, topics pertaining to education (Topic 36), neighborhood (Topic 29), and socioeconomic status (Topic 3) were still discernable from one another despite large overlap in terms associated with race and ethnicity.

**Figure 4:**
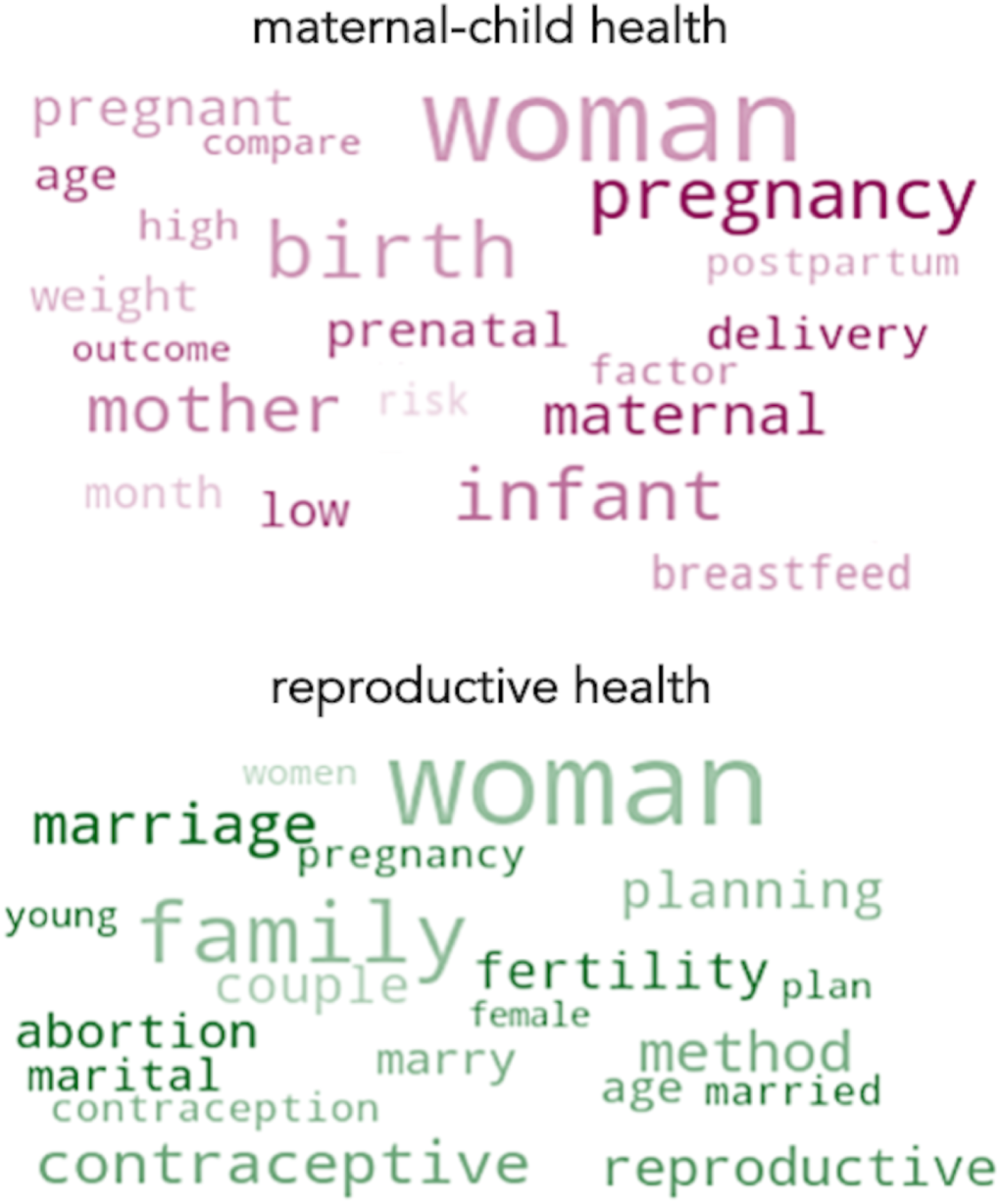
Word clouds for maternal-child health (Topic 11) and reproductive health (Topic 33). Topic modeling of HDMH article titles and abstract text enabled nuanced analysis capable of distinguishing between seemingly similar themes like maternal-child health and reproductive health despite shared terms (e.g., woman, pregnancy). Word size indicates relative frequency among topic terms. *Abbreviations*: HDMH, Health Disparities and Minority Health.

Analysis yielded 50 topics which exhibited varying levels of prevalence in the HDMH literature (**Figure 5**). We present each topic label and the percentage of articles for which that topic was the dominant theme of the article. For example, maternal-child health (Topic 11) was the dominant theme for slightly less than 3% of articles and reproductive health (Topic 33) was the dominant theme for slightly more than 1% of articles. Topics labeled miscellaneous 1 (Topic 50) and miscellaneous 2 (Topic 47) represent a small subset of publications (0.03% and 0.2% of articles, respectively) that lacked a discernably coherent theme upon manual inspection. We also present the top 5 most representative articles for each topic (**Table S2**).

**Figure 5:**
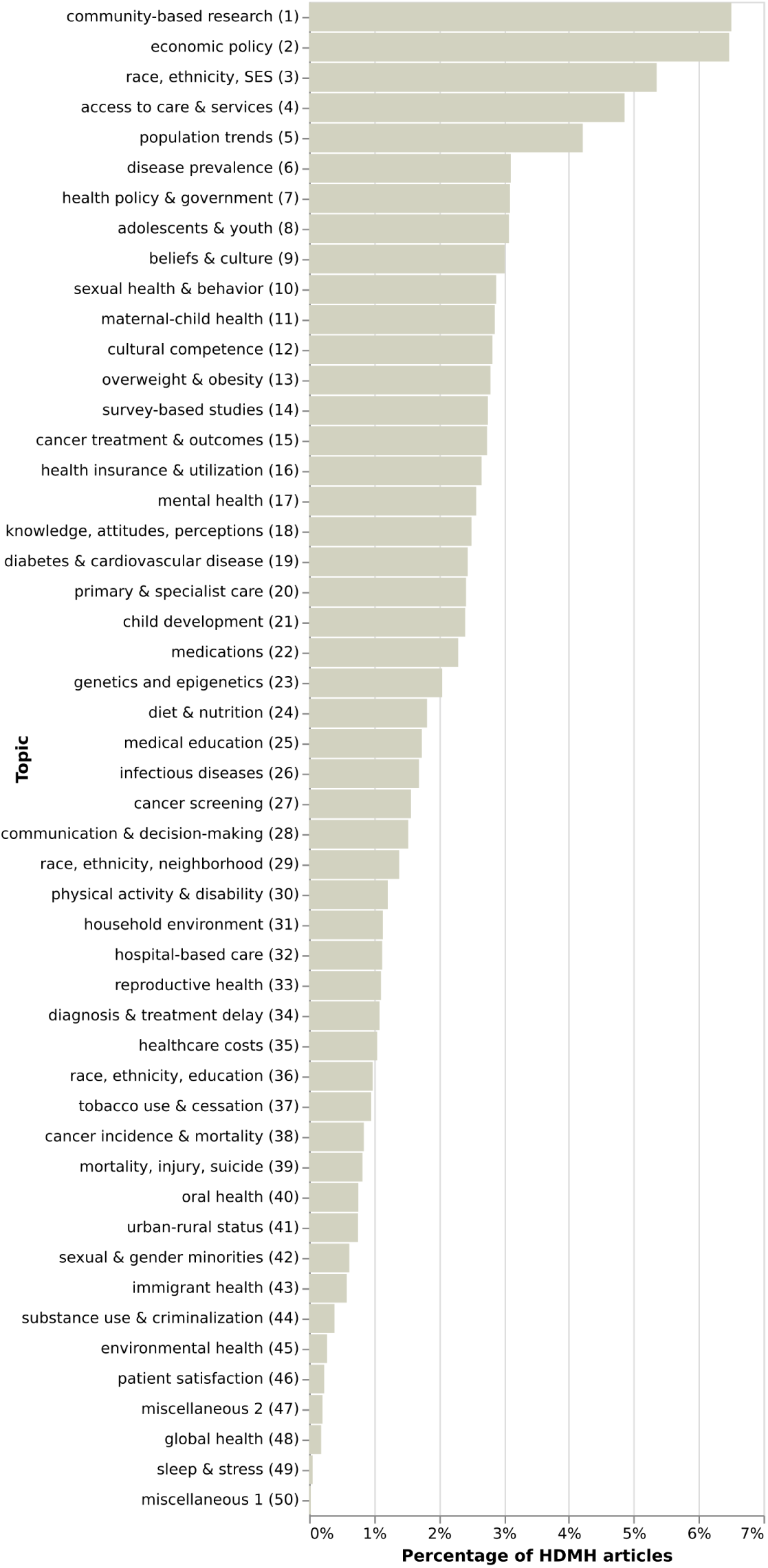
Topics generated from titles and abstracts of HDMH articles (N=206,011). Topics spanned 50 themes that ranged in discussion of populations (e.g., racial and ethnic groups, sexual and gender minorities, adolescents, immigrants), conditions (e.g., cancer, cardiovascular disease, diabetes mellitus, infectious diseases, and overweight/obesity), and study methods (e.g., community-based research, assessments of knowledge, attitudes, and perceptions). For each topic, we present the percentage of articles for which that topic was the dominant theme of the article.

Community-based research (Topic 1) is a major theme in the HDMH literature. Overall, it was the dominant topic for 6.5% of articles, the largest percentage among all topics generated (**Figure 5**). Examples of studies involving community-based research methods include dissemination of research on community health between Alaska Native villages and academics (33), increasing the relevance of research to underserved communities by engaging community health workers (34), and creating faith-based networks to conduct collective health assessments (35).

Economic policy (Topic 2) and race, ethnicity, and socioeconomic status (Topic 3) were the second and third most common themes comprising 6% and 5% of reported studies, respectively (**Figure 5**). Articles related to socioeconomic status concerned, for instance, the family unit and household wealth (36) and the impact of income inequality on self-rated health (37), while publications on economic policy discussed responses to global economic crises (38), trends in urban employment (39), and combating chronic poverty (40) among other issues. Access to care and services (Topic 4) was the dominant theme for 5% of articles (**Figure 5**). Related studies included reports on barriers to end-of-life planning (41), poor access to allied health services (42), and lack of support for people living with HIV to meet basic, psychosocial, and other needs (43).

The least dominant theme among all articles was sleep and stress (Topic 49). Overall, this theme comprised 0.05% of the literature in total (**Figure 5**). Examples of sleep and stress studies include the impact of shift work on sleep duration and health (44), sleep patterns among gender minority adolescents (45), and the influence of race and socioeconomic status on sleep (46).

Temporal trends in topic prevalence within the HDMH literature were also evident (**Figure 6**). For example, cancer treatment and outcomes (Topic 15) exhibited the largest rise in coverage over the last half-decade with a substantial increase from 0.3% of HDMH articles published in 1975 to 5.7% in 2020 (**Figure 6A**). Reporting on cancer screening (Topic 27) also grew from 0.3% in 1975 to a peak in publication output of 2̃% starting in the mid-to-late 1990s (**Figure 6A**). In contrast, cancer incidence and mortality (Topic 38) remained relatively stable as the dominant theme for 1% of all HDMH articles each year (**Figure 6A**).

**Figure 6:**
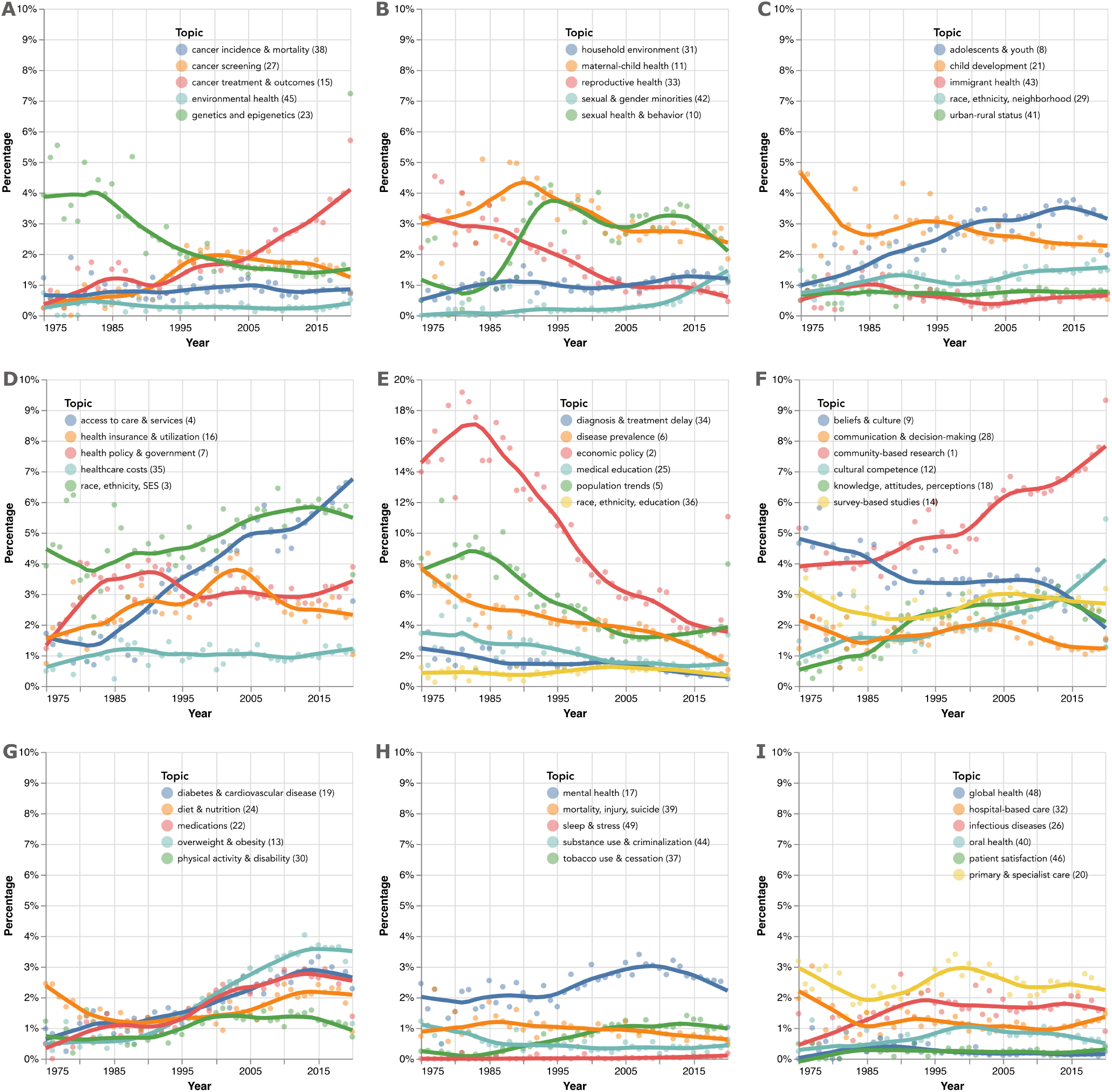
Topics generated from the health disparities and minority health bibliome (206,011 articles) by publication year, 1975-2020. Trend lines representing annual percentage of articles based on the dominant topic of each article are graphed using LOcally Estimated Scatterplot Smoothing (LOESS). Themes in the literature have changed over the last half century, ranging in discussions of groups and subgroups (e.g., racial and ethnic groups, sexual and gender minorities, adolescents, immigrants), conditions (e.g., cancer, cardiovascular disease, diabetes mellitus, infectious diseases, and overweight/obesity), and study approaches (e.g., community-based research; assessments of knowledge, attitudes, and perceptions). *Abbreviations:* SES, socioeconomic status.

During this same period, sexual and gender minorities (Topic 42) steadily emerged as a nascent theme in the literature (**Figure 6B**). Topic prevalence among HDMH articles rose annually from 0% in 1975 to 1.3% of articles by 2020. Sexual health and behavior (Topic 10) also saw a net increase in prevalence (0.5% to 1.1% of HDMH articles from 1975 to 2020, respectively), though peak annual coverage occurred in the early 1990s (**Figure 6B**). maternal-child health (Topic 11) and reproductive health (Topic 33) represent a consistent percentage (3̃% and 1̃%, respectively) of HDMH articles published over the last fifteen years (**Figure 6B**). This period of stability, however, follows a drop from much higher coverage during the 1980s (3-5% of articles annually).

Publication trends among other population groups were also noteworthy. For example, studies regarding adolescents and youth (Topic 8) increased from 1.0% in 1975 to 2.0% in 2020, while articles specifically focused on child development (Topic 21) have fallen from 4.7% to 0.5% in that same time frame (**Figure 6C**). By comparison, articles reporting on immigrant health (Topic 43) encompassed roughly 1% of the HDMH literature from year to year (**Figure 6C**).

Studies involving race, ethnicity, and socioeconomic status (Topic 3) comprise a fairly consistent percentage of the literature each year as well with an annual prevalence between 5% and 6% over the last two decades (**Figure 6D**). HDMH articles concerning healthcare costs (Topic 35) and health policy and government (Topic 7) also appear relatively stable at nearly 1% and 3%, respectively, over the last 30 years (**Figure 6D**). In contrast, discussion of access to care and services (Topic 4) saw marked growth from 1.7% of HDMH articles published in 1975 to as high as 6.6% in 2019, while studies concerning health insurance and utilization (Topic 16) decreased from a peak of nearly 4% in the late 1990s to 2-3% annually over the last ten years (**Figure 6D**). Most notably, HDMH studies focusing primarily on economic policy (Topic 2) declined precipitously from 19% of articles produced in 1981 to 4.3% in 2019 (**Figure 6E**).

Traditional epidemiologic studies concerning population trends (Topic 5) and disease prevalence (Topic 6) also fell in popularity, with prevalence for each dropping from 8% in 1975 to 3.9% and 2.0%, respectively, by 2019 (**Figure 6E**). Similarly, articles regarding diagnosis and treatment delay (Topic 34) decreased from 2.5% to 0.5% during this same period (**Figure 6E**). Studies concerning medical education (Topic 25) also saw a decline, falling from 3.4% to 1.5%, though more general reporting on race, ethnicity, and education (Topic 36) was stable with a mean prevalence of 1.0% during the last half-decade (**Figure 6E**).

Community-based research (Topic 1), in contrast, demonstrated a surge in prevalence, nearly doubling from 5.2% in 1975 to 9.3% in 2020 (**Figure 6F**). Notably, research on cultural competence (Topic 12) rose during this time from 1.5% of the literature to 5.5% as well (**Figure 6F**). Reporting on general beliefs and culture (Topic 9), however, declined by nearly half from 4.7% in 1975 to 1.8% in 2020, while assessments of knowledge, attitudes, and perceptions (Topic 18) quadrupled from 0.8% in 1975 to 3.3% of articles annually by 2013 (**Figure 6F**). Studies focusing on communication and decision-making (Topic 28) routinely comprised 1-2% of HDMH articles produced annually (**Figure 6F**).

Related areas of research such as overweight and obesity (Topic 13) and diabetes and cardiovascular disease (Topic 19) exhibited comparable patterns of growth initially ranging in prevalence from 0.5% to 1.0% in 1975 and more recently comprising 3.3% to 3.7% in 2015 (**Figure 6G**). During this same time, literature on physical activity and disability (Topic 30) maintained a modest yet relatively stable prevalence of 1̃% (**Figure 6G**). Reporting on diet and nutrition (Topic 24), however, has cycled between an initial annual prevalence of 2.5% in 1975 to a low of 1.0% in 1985 and returned to 2.3% again by 2017 (**Figure 6G**).

In contrast, reporting on mental health (Topic 17) has not changed drastically over the last half-decade, remaining at approximately 2-3% of articles since 1975 (**Figure 6H**). Similarly, reporting on substance use and criminalization (Topic 44) has been consistent, comprising on average 0.5% of articles published each year since the 1980s (**Figure 6H**). Studies regarding tobacco use and cessation (Topic 37) have seen slight growth over the years with a prevalence of 0.3% in 1975 and 1.0% in 2019 (**Figure 6H**). By comparison, articles focusing on infectious disease (Topic 26) rose from a low of 0.5% in 1976 to a high of 2.8% in 1990 and maintained an annual prevalence of 2̃% through 2020 (**Figure 6I**).

### 2.6 HDMH Monitor, a new resource for discovery

To support further navigation of the HDMH bibliome, we developed HDMH Monitor, a publicly available interactive dashboard with an accompanying article repository (https://hdmhmonitor.dbmi.columbia.edu). Functionality includes display of summary statistics and data visualizations in addition to the ability to search and copy, print, or download article metadata for further study and analysis. Each article entry in the repository is indexed by a number of elements including condition chapter and disease category code(s), population(s) of interest, study type, and dominant topic, among others.

## 3. DISCUSSION

Leveraging computational methods and open-source data enabled a large-scale scoping review of the HDMH bibliome. Our comprehensive examination of the literature identified 206,011 HDMH studies published in MEDLINE between 1975 and 2020. Publications concerning HDMH have increased in recent years yet represent only a small fraction (1̃%) of the body of literature found in MEDLINE. Indexing articles based on a combination of MeSH, disease codes, and latent topics allowed for complementary analyses and findings regarding dominant study methods, (sub)populations, conditions, and themes in the HDMH literature. By extension, this also allowed for elucidation of less well studied domains in the HDMH literature to which clinicians, public health practitioners, policy makers, and researchers might dedicate attention in the future.

### 3.1 Informing Opportunities for Extending HDMH Research to a Wider Range of Conditions

Processing text from more than 200,000 articles revealed that half of all HDMH studies in MEDLINE report on just three disease areas (cancer, mental health, and endocrine disorders), while nearly a quarter of articles mentioned either external causes of morbidity and mortality or consequences of external causes (e.g., injury or poisoning). Even if these condition categories genuinely represent the greatest sources of disparity or disease burden, the relative dearth of studies regarding other conditions and opportunity costs associated with such concentrated efforts cannot go unappreciated. Our gap analysis suggests that perhaps there are indeed disparities in the disparities literature and offers potential areas that may merit additional study (for example, dermatologic, vision, and hearing-related needs of minority populations). While there are numerous factors that result in concentrated study in particular research areas, low levels of dedicated funding to examine inequities associated with these conditions and a dearth of specialists from diverse backgrounds are both possible contributors to reduced attention.

As a preliminary attempt at fostering linkages between existing and emerging data sources, we sought to juxtapose conditions mentioned in the literature with their relative prevalence among more than 42 million Americans using a shared terminology (ICD-10) and large claims databases stratified by insurance status (private, Medicaid, Medicare). Unsurprisingly, the prevalence of diseases in the general population did not necessarily reflect their distribution among HDMH studies. We readily concede that this crude comparison is imperfect, especially given the very definition of health disparity and our inability to stratify insurance claims by minority populations due to data constraints, but our approach also demonstrates that comprehensive assessment and comparison of potential conditions is feasible. The ICD-10 terminology comprises more than 70,000 diseases, signs and symptoms, abnormal findings, complaints, social circumstances, and external causes of injury or diseases. The hierarchical coding structure of ICD enables summation at varying levels of granularity and allowed us to evaluate many conditions at a high, digestible level appropriate for a scoping review. In identifying conditions present in the literature, by extension, we were also able to isolate conditions that were absent in HDMH research.

### 3.2 Informing Opportunities for Expanding HDMH Research to More Diverse Populations

Our analyses also found high variability, with respect to both the overall number of articles and their distribution among conditions, when stratified by race, ethnicity, immigrant status, sexual orientation and gender identity, disability status, and rural-urban residence. Disparity in the distribution of HDMH articles concerning different minority communities is to be expected. As one might discern from the title of the Report of the Secretary’s Task Force on Black and Minority Health (i.e., the Heckler Report), a large focus of HDMH research concerns the health and well-being of Black Americans. Truly, the long-lasting multi-generational impacts of slavery, Jim Crow, and other forms of institutionalized and personally-mediated racism cannot be overemphasized as contributors to health disparities (47–49), nor can we ignore the tireless efforts of those who helped shape the foundation of HDMH research, much of which is based on work conducted by and devoted to improving conditions for Black Americans. Current patterns of demographic disparity related to the COVID-19 pandemic only serve to illustrate the persistence of disproportionate burden of disease and premature mortality experienced (32).

### 3.3 Study Designs in the HDMH Literature

The MeSH analysis elucidated trends in study designs. Notably, reporting on RCTs remains quite low (4% of the HDMH corpus). Achieving meaningful, lasting reductions in health disparities will likely prove elusive without rigorously evaluated interventions – the third and final pillar of the NIMHD science visioning plan. Fortunately, we found that the annual number of articles reporting on RCTs increased five-fold over the last two decades illustrating that intervention-based research is on the rise.

### 3.4 Historical Analysis Reveals the Close Link between National Initiatives and Research Areas of Focus

Historical patterns were also explored via topic modeling, a quantitative analog to the qualitative method of iterative thematic analysis, which revealed decline and emergence of new themes in HDMH literature over the last half century. This approach offered complementary information and allowed for a nuanced analysis. Summarization based on ICD chapter alone would have confounded differences in trends related to maternal-child health and reproductive health, for example. Similarly, cancer incidence and mortality, cancer treatment and outcomes, and cancer screening were three separate topics that would have been subsumed by a single ICD chapter.

Trend analysis illustrates the potential impact of attention and funding on HDMH research. Examination of HDMH publications in MEDLINE saw increased output immediately after the Heckler Report was released in 1985 and then again in 2000 when the National Center on Minority Health and Health Disparities (NCMHD) was formally established (later redesignated as the NIMHD) by Congress. As a more concrete example, the emergence of a sizable body of literature regarding sexual and gender minorities starting in the early 2000s aligns very closely with increased NIH funding for sexual and gender minority health, in particular, funding not solely tied to HIV/AIDS-related research (50).

### 3.5 A Systematic, Scalable, and Reproducible Approach for the Scoping Review

Given the sheer size of the corpus of literature, a traditional manual review approach would not have been practical. Methods were needed to streamline a standardized, scalable, and reproducible review process. Employing existing ontologies and a closed-world assumption (i.e., all possible conditions are represented in those terminologies) facilitated top-down assessment of studied conditions and, by extension, those conditions not well represented within and across populations. As a complement, topic modeling allowed for a bottom-up identification of emerging themes based solely on the text. Further, using common terminology (i.e., ICD-10) for both biomedical literature and insurance claims data allowed for direct comparison across data sources. It is likely reasonable to assume that our computational approach to performing a scoping review is generalizable for use on any large body of literature, regardless of domain.

Lastly, as scalable, reproducible methods are needed to process, characterize, and monitor trends in HDMH research systematically, we developed HDMH Monitor, a new resource that builds on our scoping review. To improve utility and reduce barriers for further study of specific clinical conditions or major themes, we expanded traditional MEDLINE indexing to include ICD-10 chapter and disease category codes as well as topics. By automating search and archive of HDMH articles, we seek to enable investigators, clinicians, and policy makers alike to quickly navigate and explore the HDMH bibliome. In doing so, we hope to support further discovery and continued comprehensive examination of the literature, so that health disparities findings may be more swiftly translated into health disparities reductions. We hope that this work will facilitate hypothesis generation, spur future investigation, and provide a tool for benchmarking progress in advancing health equity across the myriad intersectional dimensions of individual and group identity.

### 3.6 Limitations

We recognize that our study has certain limitations. Most notably, our examination of the literature was limited to a single information source, MEDLINE. As the primary tool for performing biomedical literature searches, this was an obvious choice and had the added advantage of enabling us to employ an existing query strategy developed by the NLM. While this allowed for a robust and validated approach, the strategy was last updated in 2019 (but can still be used to retrieve current publications), so may not produce the same performance if changes are made to MEDLINE indexing. Our large-scale review also incorporated a substantial number of articles and thus it was impractical to perform individual review of each article. Accordingly, while we included all articles in our analysis, we only manually reviewed a sample of articles. Our indexing approach also had potential constraints. MeSH may not have completely captured populations and study methods. Moreover, our disease mapping approach may have possibly missed conditions mentioned in the literature. Given that our topic model will require updating over time to allow for discovery of newer themes, this limits reproducibility over time. Lastly, we acknowledge that the claims dataset analysis does not necessarily represent the true prevalence in the population. Prior reports in the literature have utilized insurance claims to estimate disease prevalence with some success (51–53), but undoubtedly differential access to care and services (a major theme identified in our analysis) would likely contribute to different estimates among populations based on who actually received care as opposed to those in need of care. Conceivably, our approach can be applied to alternative data sources that may more accurately reflect true disease prevalence across diverse populations.

### 3.7 Conclusions

Computational methods were able to identify temporal trends among conditions, populations, and methods of analysis in the HDMH literature in a scalable fashion. Our comprehensive approach suggests that despite the increase in the publications in health disparities and minority health, some topics, conditions, and populations are less well represented.

### 4. METHODS

### 4.1 Study Design

Guided by the NIMHD research framework, we based our study design on the Preferred Reporting Items for Systematic review and Meta-Analyses extension for Scoping Reviews (PRISMA-ScR) and adapted several computational approaches to conduct our scoping review and develop our HMDH dashboard and article repository, including elements pertaining search, data charting (i.e., extraction), and synthesis of results (54).

### 4.2 Defining Health Disparity and Minority Health

The definitions of health disparity and minority health have evolved over the years. Initially, the two terms were largely conflated given that, for many conditions, disparities in health among minority and non-minority populations in the United States were commonly assumed. We based our work on current NIMHD definitions – a health disparity is a “health difference, on the basis of one or more health outcomes [e.g., higher incidence and/or prevalence and earlier onset of disease], that adversely affects disadvantaged populations” while minority health pertains to the “health characteristics and attributes of racial and/or ethnic minority groups defined by the Office of Management and Budget (OMB), who are socially disadvantaged due in part by being subject to potential discriminatory acts” (55).

### 4.3 Research Framework

Our work was informed by the NIMHD research framework, a multilevel, multidomain model that articulates a broad set of health determinants relevant to understanding and addressing minority health and health disparities and promoting health equity (55). The framework itself blends two preexisting models: the National Institute on Aging (NIA) health disparities research framework (which analyzes health disparities based on several domains including the biological, behavioral, sociocultural, and environmental) and the socioecological model (which contends that health and human development are influenced by factors spanning the individual to the macro or societal level). In the NIMHD research framework, health disparity populations are based on race, ethnicity, socioeconomic status, rural vs urban status, and sexual or gender minority status. It also recognizes other fundamental characteristics such as sex and gender, disability status, and geographic region of residence. Prior work has demonstrated that a theoretical model or framework can serve to benefit the process of evidence synthesis, facilitate connection of key concepts, direct search strategy, structure and clarify results, help identify knowledge gaps, and reveal areas for future research (56).

### 4.4 Information Sources, Search, and Selection of Sources of Evidence

For our data source, we used the MEDLINE database, a bibliographic repository that contains more than 28 million biomedical references from over 5,200 journals. MEDLINE contains literature published from 1966 to present supplemented with selected references published prior to 1966. For our review, we restricted our search to articles published between 1975, just prior to the release of the landmark Heckler Report that brought unprecedented attention to health disparities, and 2020 in light of the recent arrival of the COVID-19 pandemic. MEDLINE and its public search interface, PubMed, are the primary tools for performing biomedical literature searches among health professionals and students (25–28).

Using this data source also enabled us to employ the MEDLINE/PubMed Health Disparities and Minority Health Search Strategy established by the NLM (24). This strategy incorporates 257 subject terms and free-text strings found in titles and abstracts. We included all articles published in English regardless of study design or article type. To minimize selection bias, our search only included articles with publication dates between 1975 and 2020, inclusive, and did not incorporate any disease-related terms. We also limited our search to articles with abstracts containing a minimum of fifty words to ensure sufficient text for information extraction and characterization.

Additionally, we incorporated information from electronic health record and medical claims data sources to estimate disease prevalence using records from approximately 42 million Americans using the MarketScan Commercial Claims And Encounters (CCAE), Medicaid (MDCD), and Medicare (MDCR) datasets.

### 4.5 Data Charting Process and Data Items

On 25 August 2021, we queried the MEDLINE database via its public application programming interface (API) using the parameters described above. For each article, we extracted the PubMed reference number (PMID), publication date, title, abstract, language, and Medical Subject Headings (MeSH) terms using the open-source Entrez package of the Biopython Python library. Once all abstracts and article metadata were retrieved, we removed records for any articles containing fewer than fifty words in their abstracts in light of eligibility criteria noted above. We also performed a general query of MEDLINE to obtain the total number of English articles published each year using similar restrictions as described above but excluded parameters from the HDMH search strategy.

Briefly, to identify trends in diseases, populations, and methods of analyses in the HDMH literature, we indexed articles according to (1) their MeSH terms, (2) conditions mentioned in titles and abstracts, and (3) latent topics generated from articles using a form of unsupervised ML called probabilistic topic modeling. Leveraging our clinical data sources, we also estimated the prevalence of conditions based on the ICD-10 concepts, which capture over 70,000 diseases, signs and symptoms, abnormal findings, complaints, social circumstances, and external causes of injury or diseases (57, 58).

### 4.6 Medical Subject Heading (MeSH) terms

MeSH terms are readily available and often used to characterize MEDLINE literature. Nonetheless, there is also a high degree of variability with respect to the number of MeSH terms assigned to a given article (a minimum of at least two terms), which may result in incomplete capture of the most salient thematic elements. Moreover, MeSH may lack newer themes if terms have not been added to the vocabulary yet. In this sense, the inclusion of MeSH in our analysis is logical and necessary but not sufficient. We utilized MeSH terms to identify populations of interest (e.g., Black or African American descent, immigrant status, or rural residence) and categorize articles according to their study design (e.g., cohort study or randomized controlled trial).

### 4.7 Biomedical Named-entity Recognition (NER)

Prior work has demonstrated that MeSH terms do not fully map to all ICD-10 concepts. Given its comprehensiveness, we chose to index by ICD-10 as it lends itself to a systematic and scalable assessment of the vast majority of, if not all, potential clinical conditions that might be represented in the health disparities and minority health literature. Additionally, use of ICD-10 lends itself to compare statistics regarding conditions across seemingly disparate data sources (e.g., biomedical literature and claims databases).

We indexed conditions mentioned in titles and abstracts through NER using MetaMap, an open-source biomedical literature tool designed and maintained by the NLM for recognizing Unified Medical Language System (UMLS) concepts via NLP (59). Specifically, we identified conditions associated with the UMLS disorder semantic types: acquired abnormality, anatomical abnormality, bacterium, congenital abnormality, cell or molecular dysfunction, disease or syndrome, experimental model of disease, finding, injury or poisoning, mental or behavioral dysfunction, pathologic function, and sign or symptom. MetaMap produced UMLS concept unique identifiers (CUIs) which were then converted to their corresponding ICD-10 codes using the UMLS Metathesaurus, a compendium that maps CUIs to codes from numerous controlled vocabularies in biomedicine.

### 4.8 Probabilistic Topic Modeling

To identify latent topics in the literature, we performed a form of probabilistic topic modeling called latent Dirichlet allocation (LDA) using the open-source Gensim Python library (58–60). Probabilistic topic models are a class of generative statistical models capable of summarizing a large collection of documents based on a smaller number of word distributions. Detection of distributions of words (i.e., topics) within the literature, broadly speaking, operates as a scalable quantitative analog to traditional qualitative approaches of thematic analysis. Probabilistic topic modeling is an established methodology used in multiple fields, such as reviewing trends in scientific articles (58, 61).

Our LDA pipeline first involved data pre-processing steps that included concatenating the title and abstract text of each article, tokenizing that text (i.e., isolating individual words), transforming each word to lowercase, then removing punctuation, stopwords (e.g., “is”, “the”), and types with fewer than 50 tokens in the corpus. We also augmented our corpus by generating bi-grams (e.g., “smoking cessation”) and tri-grams (e.g., “acute myocardial infarction”). We optimized our LDA model using a grid search based on topic number with k topics ranging from 25 to 175 and a step size of 25. We evaluated all LDA models based on coherence scores (Fig S2) and manual review (62). Two reviewers (HRN, NE) independently labeled topics generated by the optimal model based on the top 20 most representative terms within each topic (resulting in 80% agreement) and adjudicated any differences to establish consensus. After final assignment, we also performed confirmatory spot checks by reviewing the top 5 most representative articles for each topic (**Table S2**).

### 4.9 Synthesis of Results

We generated summary statistics regarding publication output over time by calculating the number of articles in MEDLINE and percentage of HDMH articles published each year of the review period. We used MeSH terms to identify study design and calculated the number of articles published in each category (e.g., randomized controlled trial) per year of the review period.

Probabilistic topic modeling identifies a mix of topics in each document. We labeled articles according to their most dominant topic based on maximum topic percent contribution. We then calculated the frequency of dominant article topic by year and graphed dominant topic prevalence rates over time using LOcally Estimated Scatterplot Smoothing (LOESS) to generate trend lines (63).

ICD-10 has a hierarchical coding structure that enables summation at varying degrees of granularity. Leveraging this hierarchy, we collapsed individual ICD-10 codes to the chapter (I-XXI) and category (A00-U89) levels for analysis. We then calculated the percentage of articles assigned codes from each chapter and category discussed in the HDMH literature as well as the prevalence of these codes in our clinical data sources. To examine the distribution of articles concerning different ICD-10 condition chapters across various populations, we generated a heatmap based on a combination of MeSH term population categories and NER-based condition tags. To determine the degree to which conditions are more or less well represented in the literature relative to their actual prevalence in the overall population, we compared the percentage of articles associated with an ICD-10 chapter to the prevalence of that chapter in our clinical databases.

## Acknowledgments

Preliminary data from this article were presented at the National Library of Medicine Informatics Training Conference on June 24, 2022 in Buffalo, NY and American Medical Informatics Association Annual Symposium on November 6, 2022 in Washington, DC.

## Author contributions

Conceptualization: H.R.N.

Methodology: H.R.N., N.E.

Investigation: H.R.N., N.E.

Visualization: H.R.N.

Supervision: N.E.

Writing—original draft: H.R.N.

Writing—review editing: H.R.N., S.B., N.E.

## Data and materials availability

Data needed to evaluate the conclusions in the paper are present in the paper and/or the Supplementary Materials.

## Appendix

**Table S1.** Top 5 most representative articles for each topic based on topic percent contribution.

| Topic | Topic Label | PubMed ID | Article Title |
| --- | --- | --- | --- |
| 1 | community-based research | 17602411 | Disseminating research on community health... |
| 1 | community-based research | 23728049 | Increasing the relevance of research to un... |
| 1 | community-based research | 28603154 | Community Building Community: The Distinct... |
| 1 | community-based research | 23727965 | Action-learning collaboratives as a platfo... |
| 1 | community-based research | 31468833 | A conceptual model for establishing collab... |
| 2 | economic policy | 17483551 | Katrina perspectives... |
| 2 | economic policy | 24319400 | Harnessing poverty alleviation to reduce t... |
| 2 | economic policy | 20737738 | Governing chronic poverty under inclusive ... |
| 2 | economic policy | 24450021 | Responding to the global economic crisis: ... |
| 2 | economic policy | 12315450 | Urban employment: research and policy in L... |
| 3 | race, ethnicity, socioeconomic status | 8211562 | Economic well-being of the old old: family... |
| 3 | race, ethnicity, socioeconomic status | 16006419 | Socioeconomic position and self-rated heal... |
| 3 | race, ethnicity, socioeconomic status | 12893616 | Income inequality, household income, and h... |
| 3 | race, ethnicity, socioeconomic status | 11820682 | Metropolitan area income inequality and se... |
| 3 | race, ethnicity, socioeconomic status | 30146496 | Association of Suicidal Ideation with Demo... |
| 4 | access to care & services | 10181676 | Provider and consumer perceptions of allie... |
| 4 | access to care & services | 24857931 | The Challenges of Providing Culturally Com... |
| 4 | access to care & services | 28664792 | End of life planning for 'hard to reach' c... |
| 4 | access to care & services | 22396929 | The NSW Refugee Health Service - improving... |
| 4 | access to care & services | 17570300 | Experiences and support needs of poverty-s... |
| 5 | population trends | 8286161 | An epidemiological survey of SIDS in the S... |
| 5 | population trends | 11446428 | Inbreeding in Gredos mountain range (Spain... |
| 5 | population trends | 9399115 | The challenge of moving from welfare to wo... |
| 5 | population trends | 12179045 | Growth of rural population: a case study o... |
| 5 | population trends | 27473140 | Trends in socioeconomic inequalities in mo... |
| 6 | disease prevalence | 21121207 | Prevalence, awareness and control of hyper... |
| 6 | disease prevalence | 22216793 | Prevalence of hypertension in a sample of ... |
| 6 | disease prevalence | 18049963 | Presbycusis among older Chinese people in ... |
| 6 | disease prevalence | 9510491 | Prevalence and risk factors of hypertensio... |
| 6 | disease prevalence | 15633712 | Prevalence of anemia in adult rural popula... |
| 7 | health policy & government | 30207901 | Mexico's Seguro Popular: Achievements and ... |
| 7 | health policy & government | 28784727 | Propping Up Indian Health Care Through Med... |
| 7 | health policy & government | 22256924 | Falling through the cracks: the hidden eco... |
| 7 | health policy & government | 32489804 | World Heart Federation Briefing on Prevent... |
| 7 | health policy & government | 10538757 | Overseas health care. Grandmothers' footst... |
| 8 | adolescents & youth | 19473772 | The influence of neighborhood disadvantage... |
| 8 | adolescents & youth | 17149030 | Ecological influences on health-promoting ... |
| 8 | adolescents & youth | 28851024 | Longitudinal Relations of Economic Hardshi... |
| 8 | adolescents & youth | 23529889 | Religious coping, posttraumatic stress, ps... |
| 8 | adolescents & youth | 23400396 | Neighborhood context and perceptions of st... |
| 9 | beliefs & culture | 15762401 | Response to Kirkpatrick (2004): differenti... |
| 9 | beliefs & culture | 16178344 | Two women with multiple sclerosis and thei... |
| 9 | beliefs & culture | 21153984 | Low-income women describe fertility-relate... |
| 9 | beliefs & culture | 12713755 | Taking a stand: using psychoanalysis to ex... |
| 9 | beliefs & culture | 15983130 | Women must endure according to their karma... |
| 10 | sexual health & behavior | 24890184 | Sexual motivation, sexual transactions and ... |
| 10 | sexual health & behavior | 24521727 | Concurrent partnerships and HIV risk among... |
| 10 | sexual health & behavior | 17159586 | Unprotected sex in regular partnerships am... |

| Topic | Topic Label | PubMed ID | Article Title |
| --- | --- | --- | --- |
| 10 | sexual health & behavior | 7960307 | Acculturation, sexual behavior, and alcoho... |
| 10 | sexual health & behavior | 17377837 | High rates of sex with men among high-risk... |
| 11 | maternal-child health | 19196854 | Factors associated with exclusive breastfe... |
| 11 | maternal-child health | 28621054 | Delivery mode and breastfeeding outcomes a... |
| 11 | maternal-child health | 26866368 | Optimal Timing of Delivery among Low-Risk ... |
| 11 | maternal-child health | 28699097 | Early Preterm Birth Across Generations Amo... |
| 11 | maternal-child health | 10711549 | Interpregnancy interval and the risk of pr... |
| 12 | cultural competence | 29894403 | Integration of Indigenous healing practice... |
| 12 | cultural competence | 28968618 | Does Doctor Know Best? Cultural competence... |
| 12 | cultural competence | 29088567 | The persistence of racially-based health c... |
| 12 | cultural competence | 29190836 | Addressing patient biases toward physician... |
| 12 | cultural competence | 29190837 | Menstrual hygiene plight of homeless women... |
| 13 | overweight & obesity | 23727501 | Achieving long-term weight maintenance in ... |
| 13 | overweight & obesity | 20885173 | A study of a culturally enhanced EatRight ... |
| 13 | overweight & obesity | 18239637 | Effect of group racial composition on weig... |
| 13 | overweight & obesity | 24027958 | Effects of social support and spirituality... |
| 13 | overweight & obesity | 21035822 | Smaller weight changes in standardized bod... |
| 14 | survey-based studies | 25877953 | Psychometric properties of Functional Asse... |
| 14 | survey-based studies | 32644927 | Validity and reliability of EQ-5D-5L among... |
| 14 | survey-based studies | 15542010 | Validity and reliability of the PDQ-39 and... |
| 14 | survey-based studies | 15892443 | Is the standard SF-12 health survey valid ... |
| 14 | survey-based studies | 24609736 | Reliability and validity of the Nijmegen C... |
| 15 | cancer treatment & outcomes | 30518497 | Demographic and facility volume related ou... |
| 15 | cancer treatment & outcomes | 28601306 | Disparities in access to care and outcomes... |
| 15 | cancer treatment & outcomes | 23415239 | Effect of insurance status on the surgical... |
| 15 | cancer treatment & outcomes | 24991720 | Disparities in receipt of radiotherapy and... |
| 15 | cancer treatment & outcomes | 30274882 | Surgical resection of lymph node positive ... |
| 16 | health insurance & utilization | 9314687 | Insurance matters for low-income adults: r... |
| 16 | health insurance & utilization | 11475546 | Gaps in health coverage among working-age ... |
| 16 | health insurance & utilization | 17354373 | Access to health insurance, barriers to ca... |
| 16 | health insurance & utilization | 9637973 | Access to care: how much difference does M... |
| 16 | health insurance & utilization | 27702956 | Access To Care Improved For People Who Gai... |
| 17 | mental health | 20070226 | Symptoms of PTSD and major depression in L... |
| 17 | mental health | 8902742 | Traumatic events and physical health in a ... |
| 17 | mental health | 645412 | Rates and risks of depressive symptoms in ... |
| 17 | mental health | 27396017 | Posttraumatic Stress Disorder and Stressfu... |
| 17 | mental health | 30486508 | Biomedical Variables and Adaptation to Dis... |
| 18 | knowledge, attitudes, perceptions | 20451639 | Factors influencing the uptake of 2009 H1N... |
| 18 | knowledge, attitudes, perceptions | 9615240 | Radon awareness and reduction campaigns fo... |
| 18 | knowledge, attitudes, perceptions | 17139233 | Predictors of human papillomavirus vaccina... |
| 18 | knowledge, attitudes, perceptions | 15911193 | AIDS knowledge and attitudes in Iran: resu... |
| 18 | knowledge, attitudes, perceptions | 27396613 | Ebola hemorrhagic fever under scope, view ... |
| 19 | diabetes & cardiovascular disease | 26603128 | Change in cardiometabolic score and incide... |
| 19 | diabetes & cardiovascular disease | 15985584 | Relative elevation in baseline leukocyte c... |
| 19 | diabetes & cardiovascular disease | 26944398 | Health- and work-related predictors of wor... |
| 19 | diabetes & cardiovascular disease | 14576369 | Effect of area-based deprivation on the se... |
| 19 | diabetes & cardiovascular disease | 29890521 | Factor VIII, Protein C and Cardiovascular ... |
| 20 | primary & specialist care | 758278 | Referral-outreach program links community ... |

| Topic | Topic Label | PubMed ID | Article Title |
| --- | --- | --- | --- |
| 20 | primary & specialist care | 22864492 | Interprofessional collaborative model for ... |
| 20 | primary & specialist care | 32443134 | Improving LGBTQ Health and Well-Being... |
| 20 | primary & specialist care | 27834232 | A New Treatment For Veterans In Need: Lega... |
| 20 | primary & specialist care | 29176094 | Providing Specialty Care for the Poor and ... |
| 21 | child development | 23360640 | Reciprocal influences between maternal lan... |
| 21 | child development | 17967109 | Education and nutritional status of orphan... |
| 21 | child development | 26566635 | Classroom Age Composition and the School R... |
| 21 | child development | 8372912 | Impact of parental separation and divorce ... |
| 21 | child development | 25321852 | Early predictors of autism in young childr... |
| 22 | medications | 22413874 | The MONET trial: week 144 analysis of the ... |
| 22 | medications | 1693738 | Comparison of felodipine and hydrochloroth... |
| 22 | medications | 26018133 | Treatment Outcomes With First-line Therapi... |
| 22 | medications | 8862216 | Assessing medication adherence by pill cou... |
| 22 | medications | 18663516 | A double-blind, randomized, placebo-contro... |
| 23 | genetics & epigenetics | 8629090 | Newborn mass screening and molecular genet... |
| 23 | genetics & epigenetics | 15906717 | G6PD Viangchan and G6PD Mediterranean are ... |
| 23 | genetics & epigenetics | 27699474 | Atlas of human diseases influenced by gene... |
| 23 | genetics & epigenetics | 32423989 | Heterologous Expression and Functional Cha... |
| 23 | genetics & epigenetics | 25443111 | Race, common genetic variation, and therap... |
| 24 | diet & nutrition | 9566994 | Fortified foods contribute one half of rec... |
| 24 | diet & nutrition | 23742909 | Consumption of added sugars among U.S. adu... |
| 24 | diet & nutrition | 28610456 | Consumer heterogeneity in sugar consumptio... |
| 24 | diet & nutrition | 30027480 | Fruit and Vegetable Intake of US Hispanics... |
| 24 | diet & nutrition | 21359162 | Characteristics of prepared food sources i... |
| 25 | medical education | 9006570 | Micmac medical student becomes role model ... |
| 25 | medical education | 6823059 | A program to recruit and educate medical s... |
| 25 | medical education | 2586298 | Responding to perceived needs of the twent... |
| 25 | medical education | 3336049 | Evaluation of Gannon-Hahnemann Program to ... |
| 25 | medical education | 11730118 | Outcomes of training pediatricians to serv... |
| 26 | infectious diseases | 29544376 | Acceptability of HIV Testing Among Jail In... |
| 26 | infectious diseases | 15494814 | Prevalence of human T-lymphotropic virus t... |
| 26 | infectious diseases | 3010700 | Predictive value of a screening test for a... |
| 26 | infectious diseases | 24521722 | Effect of nucleic acid amplification testi... |
| 26 | infectious diseases | 32762656 | HTLV screening of blood donors using chemi... |
| 27 | cancer screening | 24858861 | Cultural sensitivity and health education:... |
| 27 | cancer screening | 29168060 | The Role of Physician Recommendation in Co... |
| 27 | cancer screening | 19372282 | Pap smear screening among Asian Pacific Is... |
| 27 | cancer screening | 10187068 | Factors associated with repeat mammography... |
| 27 | cancer screening | 15894664 | Physician recommendation for papanicolaou ... |
| 28 | communication & decision-making | 3929878 | General practitioner and long term care of... |
| 28 | communication & decision-making | 10491243 | Satisfaction with methods of Spanish inter... |
| 28 | communication & decision-making | 18201406 | Recruiting patients for postgraduate medic... |
| 28 | communication & decision-making | 23910630 | A disparity of words: racial differences i... |
| 28 | communication & decision-making | 9794340 | Interpreter use and satisfaction with inte... |
| 29 | race, ethnicity, neighborhood | 29430739 | Racial disparities in kidney transplant wa... |
| 29 | race, ethnicity, neighborhood | 29715639 | Racial residential segregation and racial ... |
| 29 | race, ethnicity, neighborhood | 21697256 | Metropolitan-level racial residential segr... |
| 29 | race, ethnicity, neighborhood | 27821759 | For blacks in America, the gap in neighbor... |

| Topic | Topic Label | PubMed ID | Article Title |
| --- | --- | --- | --- |
| 29 | race, ethnicity, neighborhood | 30987098 | Disentangling Race, Poverty, and Place in ... |
| 30 | physical activity & disability | 22104979 | Time spent in physical activity and sedent... |
| 30 | physical activity & disability | 24215625 | Adult self-reported and objectively monito... |
| 30 | physical activity & disability | 22964003 | Associations between active commuting and ... |
| 30 | physical activity & disability | 15933408 | Non-fatal injuries among adults with activ... |
| 30 | physical activity & disability | 10495261 | Moderate physical activity patterns of min... |
| 31 | household environment | 32879188 | Proximity Density Assessment and Character... |
| 31 | household environment | 30642395 | Community-Led Total Sanitation and the rat... |
| 31 | household environment | 20845629 | Domestic water sourcing and the risk of di... |
| 31 | household environment | 29436345 | A Cross Sectional Study of the Association... |
| 31 | household environment | 31092211 | Bacterial contamination of drinking water ... |
| 32 | hospital-based care | 25824969 | Investigating the widely held belief that ... |
| 32 | hospital-based care | 10409007 | Assessing the extent to which hospitals ar... |
| 32 | hospital-based care | 27524767 | Indirect Referral of Orthopaedic Patients ... |
| 32 | hospital-based care | 6670578 | Diagnostic and demographic classification ... |
| 32 | hospital-based care | 28837479 | Will Rural Community Hospitals Survive?..... |
| 33 | reproductive health | 9881150 | Family planning in Isparta, Turkey... |
| 33 | reproductive health | 21933466 | Concordance between partners in desired wa... |
| 33 | reproductive health | 8262172 | Comparison of contraceptive implant adopte... |
| 33 | reproductive health | 12287023 | The effect of nuptiality status variables ... |
| 33 | reproductive health | 22891741 | Who meets their intentions to stop childbe... |
| 34 | diagnosis & treatment delay | 18808730 | Analysis of causes for late presentation o... |
| 34 | diagnosis & treatment delay | 17455418 | Status of re-registered patients for tuber... |
| 34 | diagnosis & treatment delay | 6693865 | Bipolar disorder in a low socioeconomic po... |
| 34 | diagnosis & treatment delay | 28727473 | Delay in initiation of treatment after dia... |
| 34 | diagnosis & treatment delay | 8138286 | Value of Mantoux Test in diagnosis of tube... |
| 35 | healthcare costs | 24744043 | Trends in out-of-pocket health care expend... |
| 35 | healthcare costs | 17848449 | Would greater transparency and uniformity ... |
| 35 | healthcare costs | 28109188 | Impoverishing effects of catastrophic heal... |
| 35 | healthcare costs | 19491137 | Cost sharing in Medicaid and CHIP: how doe... |
| 35 | healthcare costs | 28990452 | Do Medicare Beneficiaries Living With HIV/... |
| 36 | race, ethnicity, education | 26151232 | Impact of disability and other physical he... |
| 36 | race, ethnicity, education | 25045951 | The family-study interface and academic ou... |
| 36 | race, ethnicity, education | 26377419 | The Racial School Climate Gap: Within-Scho... |
| 36 | race, ethnicity, education | 12416323 | Ethnicity, gender, and academic self-conce... |
| 36 | race, ethnicity, education | 19364202 | Stereotypes of Latinos and Whites: do they... |
| 37 | tobacco use & cessation | 26666395 | Multiple tobacco product use among young a... |
| 37 | tobacco use & cessation | 27923866 | The association between cannabis use and m... |
| 37 | tobacco use & cessation | 23865700 | Smokers who try e-cigarettes to quit smoki... |
| 37 | tobacco use & cessation | 25180077 | Gradual reduction of cigarette consumption... |
| 37 | tobacco use & cessation | 28868760 | Cannabis use among two national samples of... |
| 38 | cancer incidence & mortality | 12627516 | Trends in prostate cancer mortality among ... |
| 38 | cancer incidence & mortality | 20031937 | Incidence of testicular cancer in the Unit... |
| 38 | cancer incidence & mortality | 29809280 | Ovarian cancer statistics, 2018.... |
| 38 | cancer incidence & mortality | 11577478 | Cancer statistics, 2001.... |
| 38 | cancer incidence & mortality | 31606852 | Changing trends in liver cancer incidence ... |
| 39 | mortality, injury, suicide | 16198967 | A study of homicidal deaths in medico-lega... |
| 39 | mortality, injury, suicide | 8655752 | Almost 19 million childhood injuries resul... |

| Topic | Topic Label | PubMed ID | Article Title |
| --- | --- | --- | --- |
| 39 | mortality, injury, suicide | 1990448 | Childhood firearms fatalities: the Metropo... |
| 39 | mortality, injury, suicide | 16884854 | Study of burn deaths in Nagpur, Central In... |
| 39 | mortality, injury, suicide | 24264507 | Coronary heart disease and stroke deaths -... |
| 40 | oral health | 26333608 | Health Information Seeking Among Rural Afr... |
| 40 | oral health | 8664091 | Needs for dental information of adolescent... |
| 40 | oral health | 2049652 | The UK Panch Gam Patidar community and den... |
| 40 | oral health | 15984179 | Perceptions of dental care need among Afri... |
| 40 | oral health | 31209445 | Socioeconomically Disadvantaged Parents an... |
| 41 | urban-rural status | 2586854 | Urbanization and breastfeeding in the Phil... |
| 41 | urban-rural status | 17167187 | Urban/rural differences in the use of phys... |
| 41 | urban-rural status | 30459052 | Individual and environmental factors assoc... |
| 41 | urban-rural status | 14964932 | Rural-urban comparisons of nursing home re... |
| 41 | urban-rural status | 28634976 | Satisfaction levels with family physician ... |
| 42 | sexual & gender minorities | 30406715 | A Comparison of Violence Victimization and... |
| 42 | sexual & gender minorities | 26831853 | Sexual Victimization and Subsequent Police... |
| 42 | sexual & gender minorities | 25864147 | Multiple early victimization experiences a... |
| 42 | sexual & gender minorities | 30632801 | Bisexual Stigma, Sexual Violence, and Sexu... |
| 42 | sexual & gender minorities | 30856062 | Out and in Harm's Way: Sexual Minority Stu... |
| 43 | immigrant health | 12159631 | Armenian migration, settlement and adjustm... |
| 43 | immigrant health | 12295042 | Undocumented workers in the labor market: ... |
| 43 | immigrant health | 12287861 | Facing changes and making choices: uninten... |
| 43 | immigrant health | 12346414 | Return and dynamics: the path of labor mig... |
| 43 | immigrant health | 15782899 | Educational selectivity in U.S. immigratio... |
| 44 | substance use and criminalization | 889417 | Decriminalization of public drunkenness: t... |
| 44 | substance use and criminalization | 30334733 | Racial/Ethnic Disparities in Antimicrobial... |
| 44 | substance use and criminalization | 32119661 | Addressing disparities in the health of pe... |
| 44 | substance use and criminalization | 24421160 | Compulsory drug detention centers in China... |
| 44 | substance use and criminalization | 18279254 | The effects of deprivation on the time spe... |
| 45 | environmental health | 10677148 | Maternal bone lead contribution to blood l... |
| 45 | environmental health | 8868555 | Lead poisoning: the implications of curren... |
| 45 | environmental health | 16262505 | Trauma and cumulative adversity in women o... |
| 45 | environmental health | 26573689 | Modeling and estimating manganese concentr... |
| 45 | environmental health | 32559243 | The intersectional effect of poverty, home... |
| 46 | patient satisfaction | 10171759 | Sitting in the lab. |
| 46 | patient satisfaction | 30020953 | A note on internet use and the 2016 U.S. p... |
| 46 | patient satisfaction | 18465407 | How should hearing screening tests be offe... |
| 46 | patient satisfaction | 15562530 | Attention HIV doctors: you're doing a good... |
| 46 | patient satisfaction | 19488849 | Internet use and the network composition o... |
| 47 | miscellaneous 2 | 18717210 | Pilot study of the breast cancer experienc... |
| 47 | miscellaneous 2 | 27779462 | Understanding breast cancer screening beha... |
| 47 | miscellaneous 2 | 25639288 | Disparities in breast cancer and african a... |
| 47 | miscellaneous 2 | 32681303 | Hispanic ethnicity as a moderator of the e... |
| 47 | miscellaneous 2 | 17895427 | I've been through something: poetic explor... |
| 48 | global health | 18471797 | Changing patterns of forest malaria among ... |
| 48 | global health | 12943974 | Pyrethroid-impregnated curtains for Chagas... |
| 48 | global health | 11454246 | Annual single-dose diethylcarbamazine plus... |
| 48 | global health | 22909096 | How does an Ethiopian dam increase malaria... |
| 48 | global health | 26473360 | Animal Reservoirs of Zoonotic Tungiasis in... |

| Topic | Topic Label | PubMed ID | Article Title |
| --- | --- | --- | --- |
| 49 | sleep & stress | 30010969 | Impact of shift work schedules on actigraph. . . |
| 49 | sleep & stress | 29555126 | The influence of psychosocial stressors an. . . |
| 49 | sleep & stress | 32949142 | Sleep among Gender Minority Adolescents. . . |
| 49 | sleep & stress | 18480189 | Influence of race and socioeconomic status. . . |
| 49 | sleep & stress | 24726571 | Predictors of shorter sleep in early child. . . |
| 50 | miscellaneous 1 | 32265705 | An Increasing Trend in the Prevalence of P. . . |
| 50 | miscellaneous 1 | 17254401 | Chilean school-age children twin registry:. . . |
| 50 | miscellaneous 1 | 19001401 | C-CAT: a computer software used to analyze. . . |
| 50 | miscellaneous 1 | 32451356 | Neonatal outcomes of extremely preterm twi. . . |
| 50 | miscellaneous 1 | 11419584 | Needle-exchange participation, effectiveness. . . |

**Figure S1.**
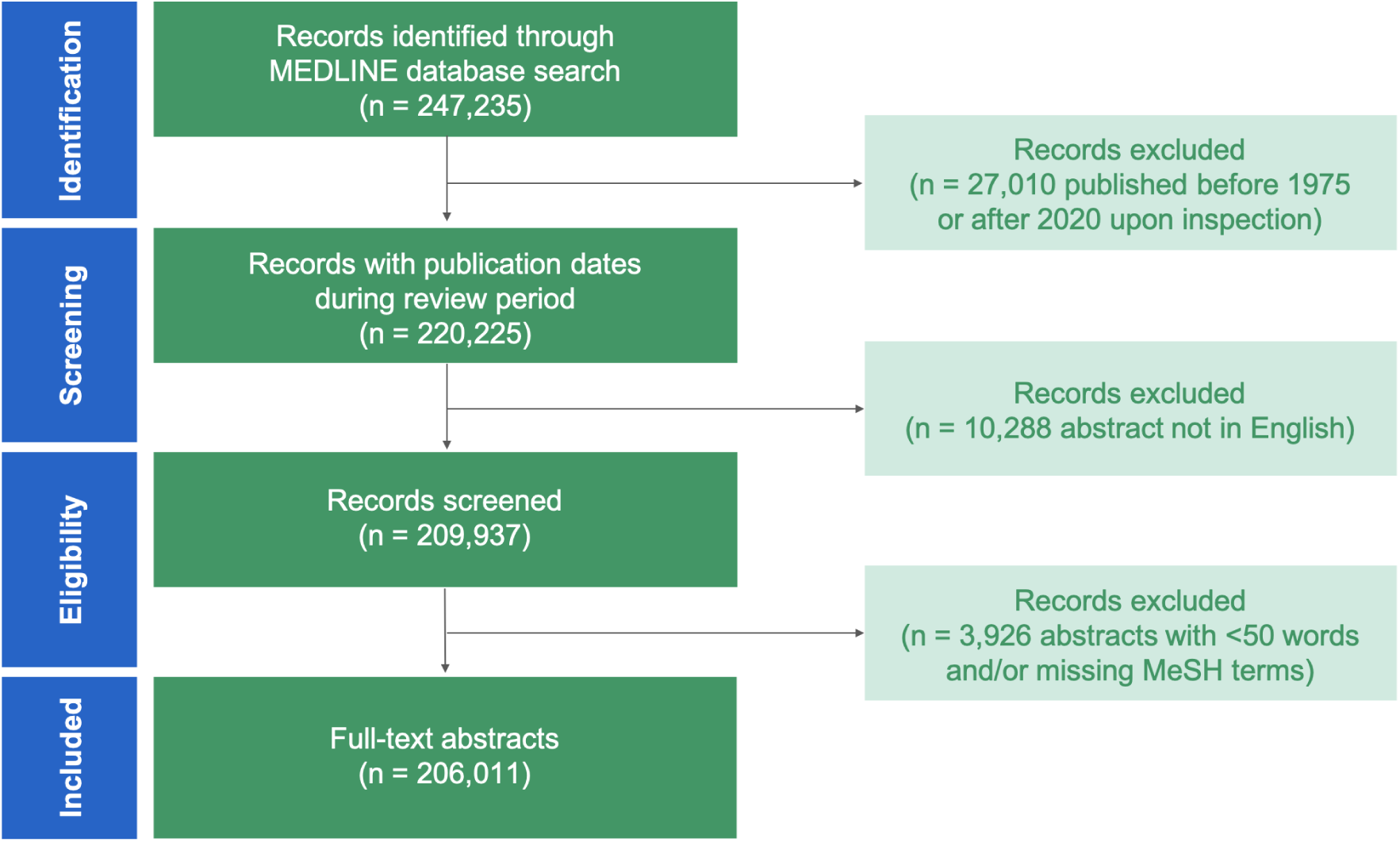
Preferred Reporting Items for Systematic reviews and Meta-Analyses extension for Scoping Reviews (PRISMA-ScR) Flow Diagram Modified for a Computational Scoping Review.

**Figure S2.**
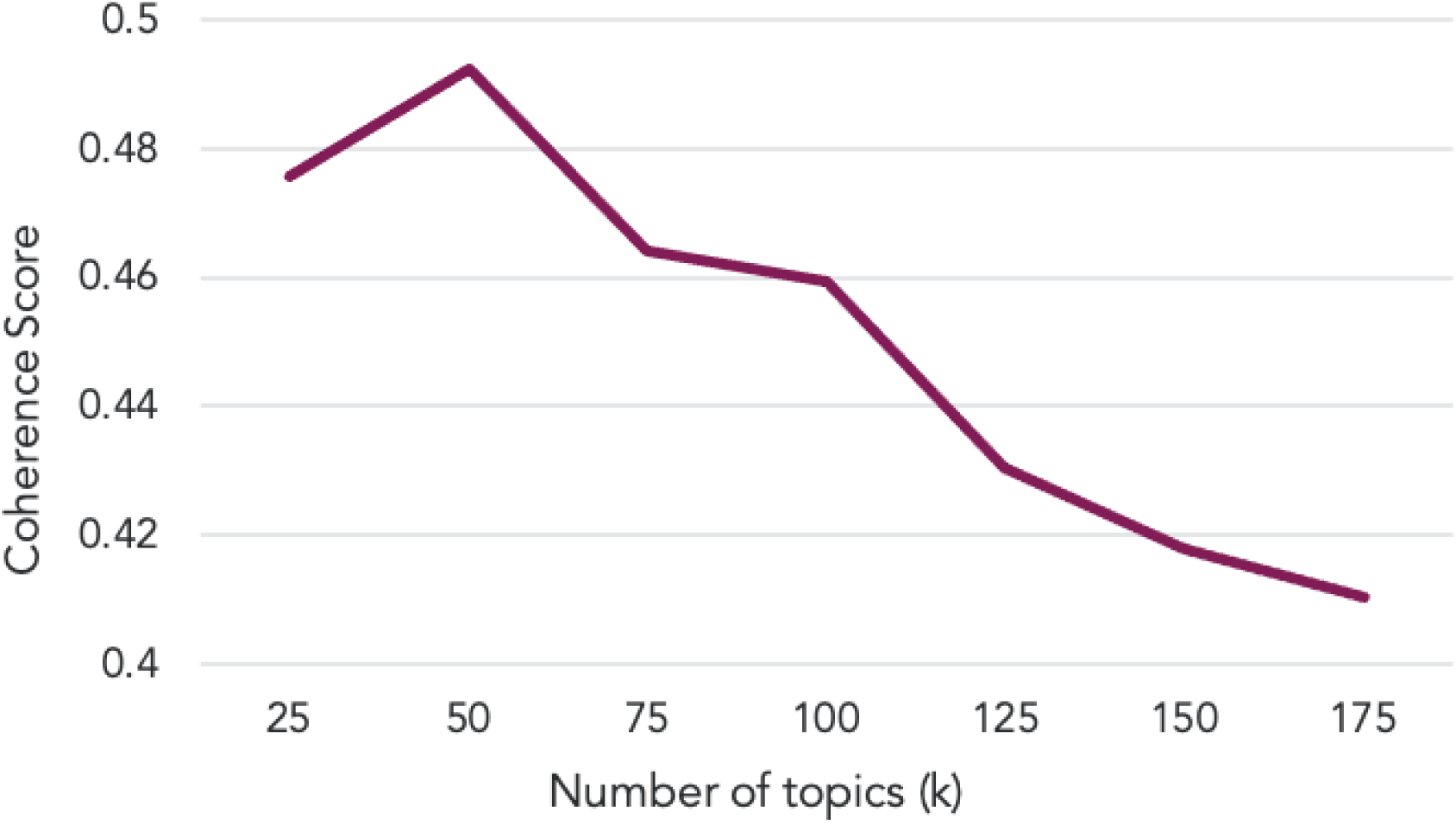
Topic model coherence (*c_v*) by number of topics (*k*), ranging from 25 to 175 with a step size of 25.

